# The utilization of contraceptives by teenagers in Lesotho: A descriptive cross-sectional survey

**DOI:** 10.1101/2024.02.09.24302554

**Authors:** Rets’elisitsoe Khiba, Isabel Nyangu

## Abstract

**Background:** Contraceptive service provision to teenagers prevents pregnancy and childbirth complications, which are the leading causes of death among teenage girls globally.

**Aim:** The study aimed to assess how teenagers utilized the contraceptive services provided at selected healthcare facilities in Lesotho.

**Methodology:** A quantitative descriptive cross-sectional design was used to collect data from 194 teenagers who were conveniently sampled from two health facilities in Berea district, Lesotho. Permission to conduct the study was sought and granted from the Ministry of Health and written informed consent was sought from the participants before they completed self-administered questionnaires. Data was captured and analyzed using the SPSS version 24.

**Results:** Most (64.5%) of the teenagers knew that contraceptives prevented pregnancy, whilst a fifth of them referred to them as the prevention of HIV or sexually transmitted infections. Utilization of contraceptives was reported by just over a third and about two-fifths were able to access them at the public health clinics daily. Injectables and condoms were the most preferred methods used by teenagers. Factors that enhanced contraceptive usage included nurses’ friendliness, as well as an understanding of why teenagers are not fully utilizing the contraceptive services provided. Most teenagers were found not to be utilizing contraceptive services due to the belief that they led to weight gain and unsatisfactory intercourse.

**Conclusion:** The study findings provided a need for and importance of contraceptive service provision to teenagers, including the need for education to improve their understanding and reduce myths.

**Implications for nursing management:** Healthcare service providers need to improve access to contraceptive services to improve their utilization by teenagers.

## 1. Introduction

About 95% of teenage pregnancies occur in developing countries, with 36.4 million women becoming mothers before the age of 18 and 5.6 million having a live birth before age 15 in 2010, Odimegwu and Mkwananzi, [5]. Hence, the use of contraceptives methods allows teenagers to postpone pregnancy, alter the timing between pregnancies, or avoid pregnancy completely. However, Munakampe, Zulu and Michelo [6] indicate that teenagers face significant barriers to contraception access and utilization that result in adverse health effects of early pregnancy and child birth. Unsafe abortions as well continue to occur partly due to failure to prevent pregnancy. Jain *et al*, [7], in their study show that teenagers need to be provided with correct knowledge for behavior change, to stop their undesirable practices and lead them to the road of healthy life.

Knowledge on contraceptives in Africa depends on the place of residence (rural/urban), religion (Christianity, Islamic, or other), and the group of peers the teenagers associate with (Ameyaw et. al,) [2]. In one study, teenagers were ignorant of contraceptive use, but when they get pregnant for the first time, the challenges they encounter raise their awareness and boost knowledge on contraceptive use including emergency contraceptives (EC), [3].

Social determinants have an influence on teenagers as teenage girls end up not using contraceptives because their boyfriends say that they make them to be wet and makes them not to enjoy sex (Proyer *et al.*) [4]. However, Nkani and Bhana, [3], pointed out that the legislation of abortion in South Africa in 1996 has seen many teenagers not worrying about getting pregnant, and hence don’t use contraceptives as they know they can have an abortion for free within 12 weeks of conception. Nevertheless, this is not the case in Lesotho, as abortion is not yet legalized, but still, they do go to South Africa to terminate the pregnancy or end up doing an unsafe abortion. Some teenage girls have difficulties in negotiating condom use because of power imbalances with their partners, which remains a barrier to successful contraceptive use, especially condoms.

Teenage pregnancy has become a challenge whereby some girl children are forced to drop out of school to raise their children. Anecdotal evidence suggests that young girls in Lesotho are seen to be running after married men who have money to buy data, so that they can communicate on social media. Unfortunately, they exchange money for airtime data on sexual intercourse and they end up falling pregnant [10]. About one-quarter of the population of Lesotho includes adolescents, therefore premarital childbearing is common and is increasing at a very shocking rate [11]. The Lesotho Demographic and Health Survey (LDHS) [12], states that 19% of women aged 15-19 had begun childbearing, while 15% had given birth already and an additional 4% were pregnant with their first child at the time of the survey.

However, even if teenagers can reach contraceptive delivery, they may not be able to get the contraceptive services they need because health service providers have knowledge gaps and misconceptions about contraceptive service provision, they do not have knowledge and skills to respond to the specific needs of teenagers or are judgemental and disrespectful to them [14]. Health providers lack the knowledge and skills to assess the cognitive, psychological, and social situation of their adolescent clients, and to offer contraception as a means of achieving their life goals by using approaches such as motivational interviewing and aspirational counselling, hence the provision of contraceptives to teenagers is low due to judgemental and disrespectful behaviour [15]. Therefore, it is significant to assess the contraceptive service provision and their utilization by teenagers.

According to Chandra-Mouli *et al.*, [8], barriers such as inaccessible service locations and cost, negatively affect adolescents and adults. However, they disproportionally affect adolescents, as they often have limited ability to move around and financial autonomy to pay for service fees and transport. Hence, in Lesotho, 19% of teenagers are seen to be pregnant by the age of 19 years old. Moreover, in Lesotho, abortion is criminalized and unsafe terminations accounts for up to 50% of inpatient deaths among females aged 13 years and older in some Lesotho hospitals [9].

### Research purpose

This study aimed to assess the utilization of contraceptive services by teenagers at two selected facilities in Lesotho.

### Research objectives

The objectives were to determine contraceptive service utilization by teenagers and describe factors that promote or hinder their use by teenagers.

## 2. Methodology

A quantitative descriptive cross-sectional design was used to collect data from 194 teenagers who were conveniently sampled and completed a self-administered questionnaire. The study was conducted at two selected hospitals in the Berea district in Lesotho. A pretest of the questionnaire was carried out on a pilot sample of 19 teenagers to assess its reliability [17]. The Cronbach’s alpha reliability coefficient was 0.8065. Data was collected over the two months of June-July 2022. As they came in for healthcare services, teenagers were introduced to the study and asked to take part. They were asked to provide voluntary written informed consent and completed the questionnaire in about 30 minutes. Participants could withdraw from the study without any prejudice towards the services they sought. Permission to conduct the study was sought from the National University of Lesotho Institutional Review Board and Lesotho Ministry of Health research and Ethics Committee (ID 45-2022) for ethical approval, prior to data collection. Data was analysed using SPSS (24) and presented using descriptive and analytic statistics.

## 3. Results

### Demographic characteristics

The majority, 73.7% (n=143), of the respondents were females, while males were making up to 23.7% (n=46), and those who were bisexual were 2.6 % (n=5). When the data were grouped, most of the respondents 83.5% (n**=**162) were teenagers who were 17 years old and above, followed by those who were 16 years old, 11.3% (n**=**22). Those who were 15 years old were 4.6% (n=9), and the least number of teenagers were those who were aged 14 or younger at 0.5 % (n=1). The mean age was 16.78. The median and mode of age was 17, while the standard deviation was 0.55. Teenagers who were in school were 60% (n=117) while 40% (n=77) of the respondents were teenagers who were not in school. Chi-square tests were done to determine the relationship between the sex (gender) of participants and having heard about the contraceptives, and it was found to be statistically significant as the P-value was below 0.05.

Teenagers who had sex 29 days and above dominated with 50.5% (n=98), followed by those who never had sex 38.1% (n=74) and those who had sex 0 to 7 days ago 8.2% (n=16). Those who had sex 21 to 28 days ago were 1.5% (n=3), as well as those who had it 8 to 14 days were 1.5% (n=3).

### Heard about contraceptives

There was 36.6% (n=71) of single teenagers who had heard about contraceptives, followed by those who were in relationships at 33.0% (n=64). Those who were married and had heard about contraceptives were 21.1% (n=41) and those who were separated, respectively, 1.5% (n=3). However, the respondents who had not heard about contraceptives and single were ranging at 2.6% (n=5), those who were married 1.5% (n=3), and those who were in a relationship 3.6% (n=7) respectively (Table 1). The chi-square tests done in this study demonstrated that the educational level of participants had a statistically significant relationship with teenagers who ever heard of contraceptives, p= 0.013

**Table 1:**
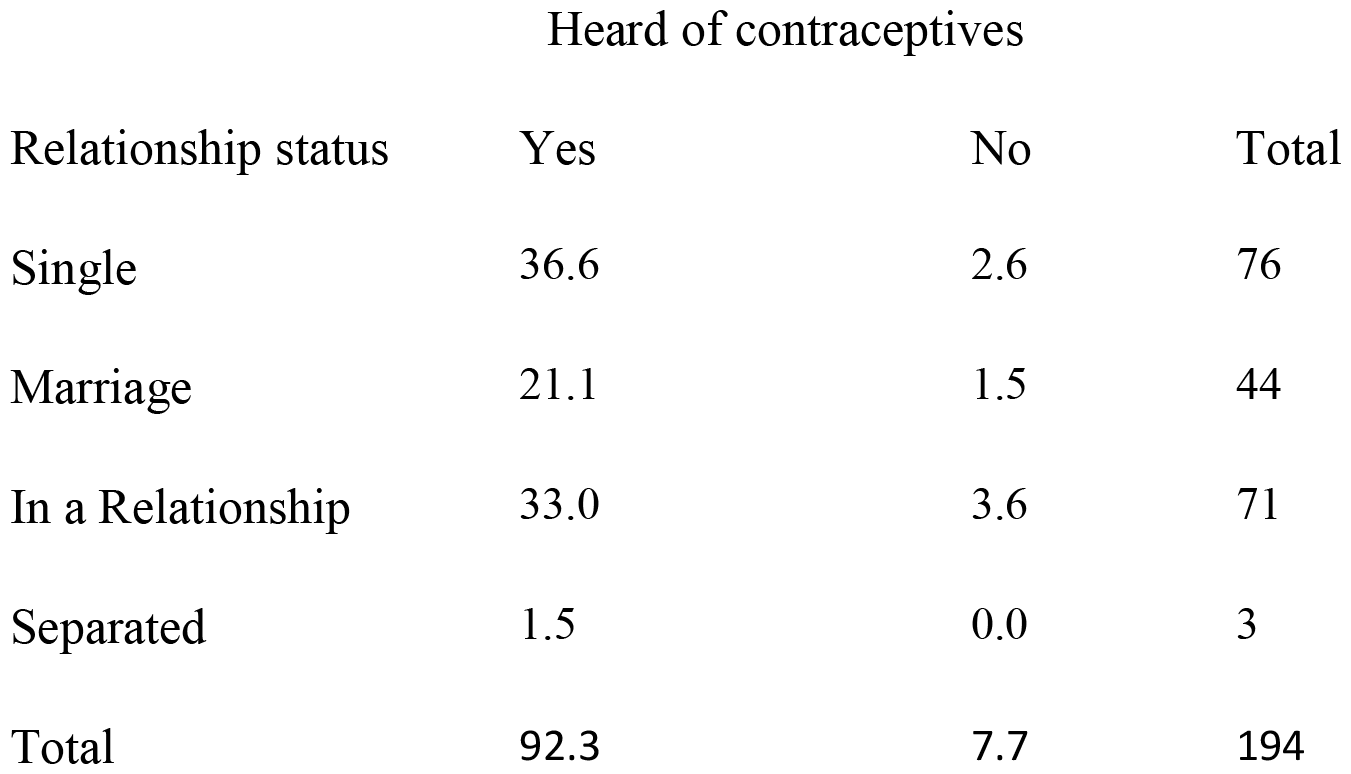
Heard about contraceptives per relationship status.

| Relationship status | Heard of contraceptives |  |  |
| --- | --- | --- | --- |
|  | Yes | No | Total |
| Single | 36.6 | 2.6 | 76 |
| Marriage | 21.1 | 1.5 | 44 |
| In a Relationship | 33.0 | 3.6 | 71 |
| Separated | 1.5 | 0.0 | 3 |
| Total | 92.3 | 7.7 | 194 |

### Understanding of the use of contraception

The findings showed that 64.2% (n=125) of teenagers perceive contraceptives as prevention of pregnancy, while 20.1% (n=39) showed that contraceptives mean prevention of HIV/STIs. However, 7.3% (n=14) indicated that contraceptive means both prevention of pregnancy & HIV/STIs, while 2.8% (n=5) responded by saying contraceptive means prolonging child spacing. Those with multiple responses were 4.5% (n=9) and the least response method of showing love & affection 1.1% (n=2). The chi-square tests were performed which determined the statistically significant association between the age of the respondents with understanding the meaning of contraception and usage of contraceptives, as well as who informed the teenagers about the contraceptives, with a p-value < 0.05. Table 2 presents the results on understanding the use of contraceptives.

**Table 2:** Understanding of the use of contraceptives.

| Description | N=194 | % |
| --- | --- | --- |
| Prevention of pregnancy | 125 | 64.2 |
| Prolonging child spacing | 5 | 2.8 |
| Showing love and affection | 2 | 1.1 |
| Prevention of HIV/STIs | 39 | 20.1 |
| Prevention of pregnancy & HIV/STIs | 14 | 7.3 |
| Multiple responses | 9 | 4.5 |
| Total | 194 |  |

### Knowledge of side effects of Contraceptives

Amenorrhea was known by 25.3% (n=49), followed by heavy menses 20.1%% (n=39) as well as weight gain and weight loss 20.1% (n=39). Only 0.5% (n=1) showed that contraceptives cause mutation, while loss of sexual drive was 10.3% (n=20). Furthermore, other respondents showed that contraceptives cause multiple effects 6.7% (n=13), however, 14.4% (n=28) did not have any idea about the side effects. Minority of teenagers responded by saying 2.6% (n=5), without being specific. Table 3 shows the results. Additionally, the chi-square tests done demonstrated that the educational level of participants had a statistically significant relationship with teenagers who heard of contraceptives (p= 0.013).

**Table 3:** Knowledge of side effects of contraceptives.

| Side effects of contraceptives | No. of Responses | % |
| --- | --- | --- |
| Amenorrhea | 49 | 25.3 |
| Causes Mutation | 1 | 0.5 |
| Heavy Menses | 39 | 20.1 |
| Loss of Sexual drive | 20 | 10.3 |
| Weight gain & weight loss | 39 | 20.1 |
| Multiple Effects | 13 | 6.7 |
| Don't know | 28 | 14.4 |
| Other | 5 | 2.6 |

### Methods that can be used to prevent unwanted pregnancy

Most teenagers prefer to use injection 38.7% (n=75) and condoms 30.4% (n=59), respectively. Oral contraceptives were 12.9% (n=25), while those who preferred multiple methods were at 14.5% (n=28). However, 3.1% (n=6) showed that abstinence is a method that can be used to prevent unwanted pregnancy, while the method was implanted 0.5% (n=1) (Table 4).

**Table 4:**
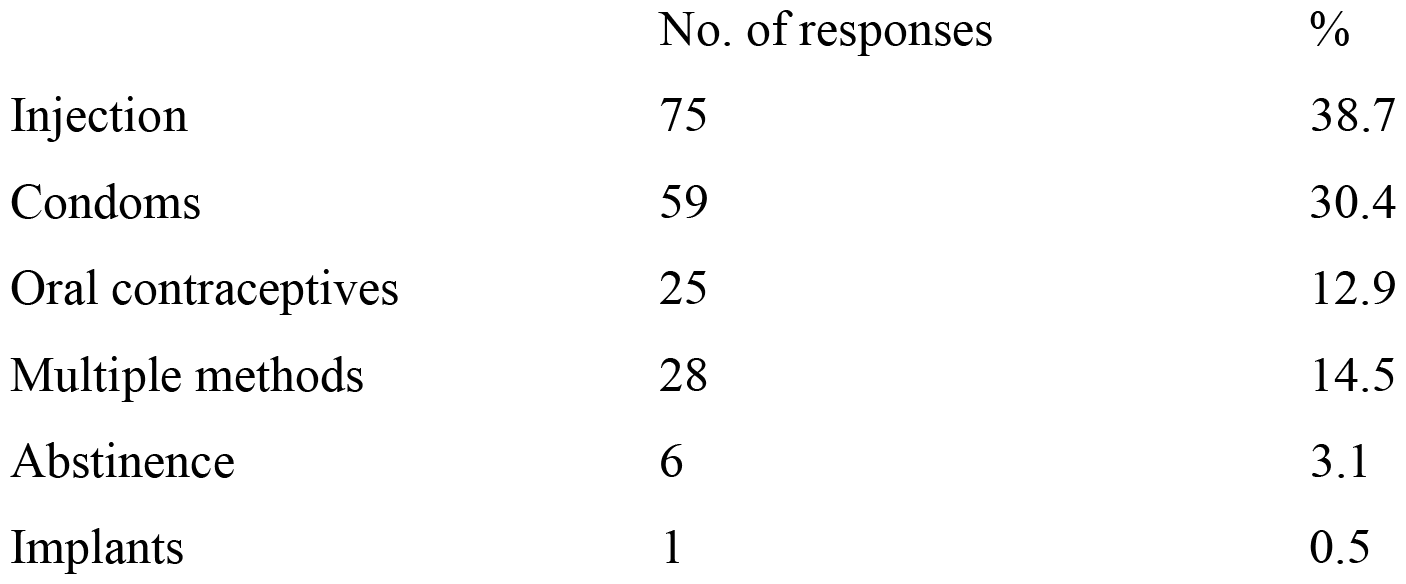
Methods that can be used to prevent unwanted pregnancy.

|  | No. of responses | % |
| --- | --- | --- |
| Injection | 75 | 38.7 |
| Condoms | 59 | 30.4 |
| Oral contraceptives | 25 | 12.9 |
| Multiple methods | 28 | 14.5 |
| Abstinence | 6 | 3.1 |
| Implants | 1 | 0.5 |

Mostly, the Catholics are the ones who have a higher percentage of not utilizing the services, however, the report shows that sometimes they utilize contraceptive services, followed by Evangelicals. The least teenagers who utilized the services were the Adventist. All in all, 26.8% (n=52) were evangelical Catholics, 39.7% (n=77), Adventist 3.6% (n=7), and Jehovah’s witness 4.6% (n=9). Those who did not belong to any religious affiliation were 3.1% (n=6), 22.2% (n=43) of those who belong to any other religion.

Thirty-two (16.5%) of the teenagers always read about contraceptives, 32.5% (n=63) read about them sometimes, while 23.2% (n=45) rarely read about them, and those who never read about contraceptives were 27.8% (n=54). As for those who talk with the health provider if they experience side effects, only 3.1% (n=6) indicated to always talk with the health provider if they experience any side effects, while 21.6% (n=42) sometimes do talk, 22.2% (n=43) rarely talk and a number of 53.1% (n=103) never talk to the health provider at all if they have side effects. 52.6% (n=102) specified that they always encourage their age mates/ friends to use contraceptives, while 18% (n=35) sometimes do that, 7.7% (n=15) rarely encourage friends, while 19.6% (n=38) have never encouraged them. Only 2.1% (n=4) showed that they do not know about the encouragement of contraceptives.

Moreover,11.3% (n=22) were respondents who always decided not to take contraceptives intentionally, 17% (n=33) sometimes, and 8.2% (n=18) rarely decided not to take contraceptives appropriately intentionally, while 57.7% (n=112) never decide to take contraceptives appropriately intentionally.5.7% (n=11) indicated that they don’t know. 58.2% (n=113) revealed that nurses always give them information about contraceptives, while 1.5% (n=3) didn’t know about nurses’ information in relation to contraceptives. 27. 3% (n=53) reported that nurses give them information sometimes, ll.3% (n=22) showed that information is given to them rarely, while only 1.5% (n=3) showed that nurses never give them information about contraceptives.

### Reasons why the best contraceptive services are not provided to teenagers

To identify the reasons why the best services are not provided, 50.6% (n=98) of respondents reported that they take too much time at the health facilities before they can access the services, while 45.6% (n=88) reported that the health providers are rude, and they have the bad attitude towards them when they must render the services. Only 3.8% (n=8) gave other reasons, like those of nurses being busy with their phones and not attending them to give the services. Figure 1 below demonstrates the information.

**Figure 1.**
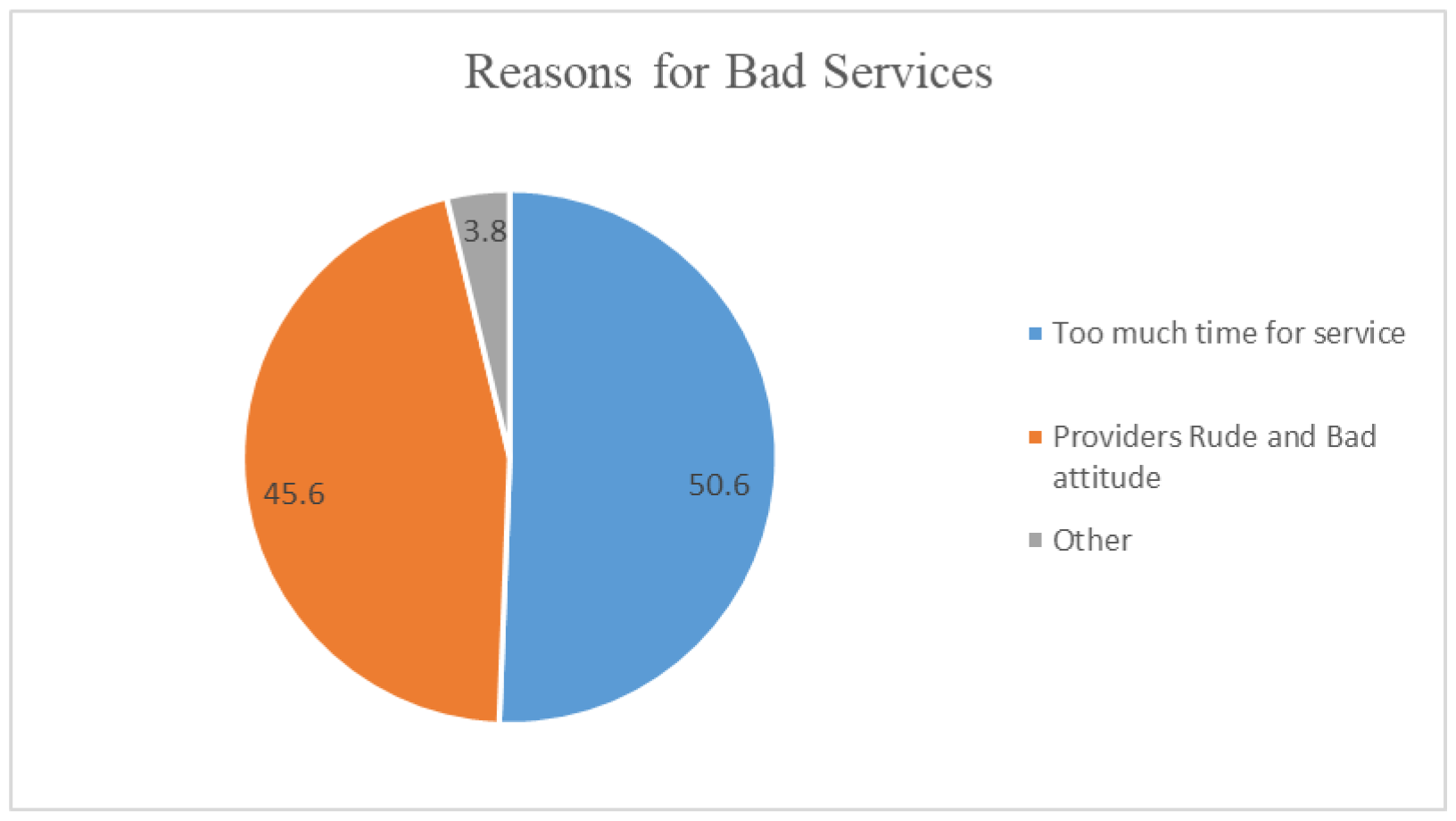
Why the best services are not provided.

## Discussion

It is significant that most of the respondents in this study were females at 73.7% while males were making up to 23.7%. However, in a study done by Woolley and Macinko [18], the adolescents who participated in the study were females at 51.7% while males were amounting to 48.3 %. Woolley and Macinko, [18] further stated that about 27.5% of teenagers were sexually initiated, and among those, approximately one in two did not use any contraceptive method during the last intercourse. This is in line with the study that was done in Nigeria by Ezenwaka et. al [19], whereby the results indicated that there was a statistically significant association between ever hearing of contraception and gender.

The findings of this study indicated that most teenagers were aged 17 years and above (83.5%), followed by those who were 16 years old (11.3%). The findings are similar to the study conducted by Young, Burke and Nic Gabhainn, [20] where they stated that as age increases, the proportion of young people reporting initiation also increases. In their study, they showed that, at 18 years old, the proportion of boys and girls reporting initiation of sex (37.3% and 54.5% respectively), are similar to the prevalence rates of sexual activity of the young adult population aged 18 to 20 years old in Ireland.

Ochen, Chi and Lawoko, [21], stated that being in school was found to be protective against teenage pregnancy, whereby periods of supervision of teenagers by teachers as well as parents could reduce opportunities for sexual activity. Additionally, contraceptive use was 24.7% among adolescents with secondary level of education [22]. However, higher pregnancies were found to be higher among adolescents who had no education. Among those who were unmarried, 8.5% used contraceptives and 8.6% of those in urban areas had used contraception.

The current study also illustrated that teenagers who had sex 29 days above dominated with 50.5%, while those who never had sex was 38.1%. Those who had sex within 0 to 7 days were ranged at 8.2%. In a study done by Young, Burke and Nic Gabhainn, [20], unprotected sex at last intercourse was reported by 10.5% of boys and 6.8% of girls.

According to Agyemang et al [23], in their study, 95% of the respondents exhibited some knowledge about contraceptives, although this high knowledge did not translate into its use as the prevalence rate was 18%. However, the present study revealed that 36.6% of single teenagers have heard about contraceptives, followed by those in relationships 33.0%, while those who were married were 21.1%. This indicates that a larger percentage have ever heard of contraceptives.

In a study done by Bakesiima *et al.*, [24], a total of 90.3% participants have ever heard of contraceptives, 67.7% had heard about modern contraceptives from a health care worker, 16.4% from family and friends, 10.9% from school and 59% from the media. A good number of participants (82.1%) knew at least two modern contraceptives and the commonly known type was the condom (70.3%).

Additionally, in a study done by Williams et al. [25], most of the respondents had heard about oral emergency contraceptive pills, and among those who were sexually active, 25.6% had reported personal use. A minority of respondents knew that emergency contraception could be bought over the counter regardless of age and gender (44.3%) and that parental consent is not required.

The most preferred method of contraception by teenagers was found to be injection (38.7%), and condoms, respectively (30.4%). Those who preferred oral contraceptives were 12.9%, while 3.1% showed that abstinence is the method that can be used to prevent unwanted pregnancy and only 0.5% would use implants. In a study done by Agyemang et al [23], condoms were mostly used contraceptives by teenagers (33%) and it was found that the perceived side effects of other contraceptives were found to be the main reason of not using contraceptives (53.7%).

Moreover, findings demonstrated that teenagers report condom use as the most common individual method of contraception [20]. Moreover, 80% of boys and girls reported using condoms at last intercourse, one-fifth of boys and one-quarter of girls reported using the contraceptive pill, and around 14% reported to be using withdrawal. About 10% of boys and 6% reported using no method of contraception at last intercourse. Additionally, Woolley and Mackinco [18] in their study showed that condom use was more widespread among both sexes, even though about one in three adolescents did not use condoms during the last intercourse.

According to Young, Burke and Nic Gabhainn, [20], in their study, findings demonstrated that teenagers report condom use as the most common individual method of contraception. Moreover, 80% of boys and girls reported using condoms at last intercourse, one-fifth of boys and one-quarter of girls reported using the contraceptive pill, and around 14% reported to be using withdrawal. About 10% of boys and 6% reported using no method of contraception at last intercourse. Additionally, Woolley and Mackinco [18] in their study showed that condom use was more widespread among both sexes, even though about one in three adolescents did not use condoms during the last intercourse.

The present study shows that mostly the Catholics are the ones who have a higher percentage of not utilizing contraceptive services, followed by the Evangelicals, however the report shows that sometimes they do utilize the services. This is in line with the study done by Ezenwaka [19] whereby it is stated that religious beliefs do constitute barriers to utilization of contraceptives among adolescents. They also stated that several religions advocate for total sexual abstinence among unmarried people and view premarital sex as immoral.

In the current study, only 52.6% agreed that contraceptives can be used by anyone, 25% were neutral, while 22.4% disagreed that they can be used by anyone. Moreover, 54.6% agreed that contraceptives are more beneficial than the side effects they give, although 31.4% disagreed, and 14% were just neutral. Those who agreed that contraceptives such as condoms prevent unwanted pregnancy were 82%, 5.7% disagreed, and only 12.3% were neutral. However, only 8.8% agreed that it is better to fall pregnant other than using contraceptives, 56.1% disagreed, and 35.1% were neutral. Additionally, 42.8% believed that contraceptives can lead to weight gain, 35.1% were neutral, while 22.1% totally disagreed that they lead to weight gain. Those who agreed that it is easy to access contraceptive services were 49.5%, 33.5% were neutral while 17% disagreed. Lastly, 55.2% agreed that school children should use contraceptives, 18.6% totally disagreed, and 26.2% were neutral about school children using contraceptives.

In another study, individual factors that limit access to contraceptives for teenagers were found to include lack of awareness and poor knowledge, fear of side effects, low self-esteem, and inability to afford the cost of services. Interpersonal barriers to access include poor- parent child communication of sexual and reproductive health matters and the negative attitudes of parents towards sexuality education of teenagers [19].

Nurse unfriendliness was found to be one of the factors that can lead to why the best services are not being rendered in the health facilities, waiting time and nurses not giving enough information. To identify the reasons why the best services are not provided, 50.6% of respondents reported that they take too much time at the health facilities before they can access the services, while 45.6% reported that the health providers are rude, and they have the bad attitude towards them when they have to render the services. Only 3.8% gave other reasons, like those of nurses being busy with their phones and not attending them to give the services.

In another study, health system barriers were found to be one of the reasons why the best services are not provided, which included lack of privacy and confidentiality, stock of contraceptive commodities, judgmental attitude of health workers, and insufficient staff that are not skilled in adolescent sexual and reproductive health [19]. Furthermore, Yakubu and Salisu [26] also stated that inadequate and unskilled health workers, long waiting time and lack of privacy at clinics, lack of comprehensive sexuality education, misconceptions about contraceptives and non-friendly adolescent reproductive services are some of the factors that contribute to why best services are not provided.

### 4.1 Recommendations

There is a need for nurses to ensure that health educations are given on the importance of using contraceptives to prevent teenage pregnancy and STIs. A trusting environment should be created for teenagers, whereby they can be able to discuss the myths and misconceptions about contraceptives, and how the use of contraceptives maybe important to them. Policy makers and program planners should make evidenced-based decisions about how to better address teenagers’ contraceptive needs. Further research is required to assess contraceptive service provision and their utilization among teenagers to produce data that is informative and useful in developing suitable interventions to address the gaps that may exist in the usage of contraceptives.

## 5. Conclusion

This study showed that the knowledge of teenagers regarding contraceptives was good. However, most teenagers were found not to be utilizing contraceptive services due to the beliefs that they had, like contraceptives leading to weight gain, while others perceived that it is not easy to access contraceptives. The study also revealed that there are factors which enhance or hinder them from accessing contraceptive services like lack of knowledge, nurses being rude, waiting time, that is, too long before they access the services. Nurse friendliness also determined whether the teenagers utilize the contraceptive services provided. The findings can help to formulate related policies that are directed at enhancing the utilization of contraceptive services.

## 6. Limitations

The study results may not be representative of the wider community of Lesotho because the teenagers who participated in the study were only those who came for service in the two selected hospitals. With the data collection tool, the questionnaire had a list of alternative responses which the respondents had to choose from, this might have created information bias. The study could be limited by the sensitivity of the nature of the questions as the use of contraceptives is associated with sexual activity. Therefore, there may be negative judgements from teenagers about contraceptives. The participants were likely to give responses that are socially acceptable instead of true responses as the questionnaires are subject to social desirability bias. The study occurred in only two institutions; therefore, the results need interpreting with caution and not generalized to the entire population of teenagers in Lesotho.

## Data Availability

Access to the data is available from the corresponding author upon reasonable request.

https://orcid.org/0000-0002-5892-9248

## Data Availability

Access to the data is available from the corresponding author upon reasonable request.

## Acknowledgements

Sincere gratitude to the teenagers who participated in the study.

